# *MAP3K7* Loss of Function Causes Dilated Cardiomyopathy

**DOI:** 10.64898/2026.09.02.26361780

**Authors:** Katherine S. Josephs, Carlos C. Smith-Diaz, Angela Woods, Claire Prince, Sean L. Zheng, Rachel Buchan, Esmé Cavanagh, Shezan Elahi, Emma Jennings, Henry Procter, James S. McTaggart, Riyad Janan, Pantazis Theotokis, Amy Baker, Natasha Henden, Kathryn A. McGurk, Katrina Prescott, Newcastle Clinical Genetics Team, Vaidehi Jobanputra, Paul James, Tina Thompson, Dominica Zentner, Upasana Tayal, Brian P. Halliday, Sanjay K. Prasad, Declan P. O’Regan, R. Thomas Lumbers, Paul J. R. Barton, Kate Richardson, Sabine Klaassen, Andres Rico-Armada, Piers E. F. Daubeney, Verity L. Hartill, Stephen P. Page, Angharad M. Roberts, David Carling, Jodie Ingles, James S. Ware, the GoDCM Consortium

**Author notes:** Correspondence (Carlos C. Smith-Diaz), (James S. Ware).

## Abstract

**Background and Aims:** Dilated cardiomyopathy (DCM) is a genetically heterogeneous cause of heart failure and sudden cardiac death. Many patients remain without a molecular diagnosis. *MAP3K7* encodes TAK1, a serine/threonine kinase important for cardiac homeostasis*. MAP3K7* variants are an established cause of syndromic disease, including cardiospondylocarpofacial syndrome (CSCF), in which DCM has been occasionally reported. Here, we demonstrate that *MAP3K7* variants can cause apparently isolated DCM, expanding the phenotypic spectrum of *MAP3K7*-related disorders.

**Methods:** We integrated four lines of genetic evidence: de novo variation, common variant association, familial segregation, and rare variant enrichment, alongside functional characterisation.

**Results:** In 117 paediatric cardiomyopathy trios from the 100,000 Genomes Project, two probands harboured rare *de novo MAP3K7* missense variants, significantly more than expected (Bonferroni-adjusted p=0.036). Independent GWAS implicated *MAP3K7* as a susceptibility locus for adult DCM. Across global DCM cohorts, we identified families harbouring rare *MAP3K7* variants, including one with segregation in nine affected relatives (p.(Tyr125Cys), estimated LOD = 2.7). Rare damaging non-truncating variants were enriched in DCM versus INTERVAL (OR 22, P=0.007) and gnomAD controls (OR 14, P=6.2×10⁻⁴), while truncating variants were associated with DCM in Genomics England and increased left ventricular volumes in UK Biobank. DCM-associated variants reduced TAK1 kinase activity, supporting a loss-of-function mechanism consistent with CSCF-associated alleles.

**Conclusion:** Multiple independent lines of evidence establish an association between *MAP3K7* loss of function variants and DCM. Several affected individuals lacked overt syndromic features, demonstrating that *MAP3K7*-related disease may present as apparently isolated DCM across the lifespan and supporting inclusion of *MAP3K7* in DCM diagnostic pipelines.

**GRAPHICAL ABSTRACT:** *Key Question: Do *MAP3K7* variants cause isolated dilated cardiomyopathy?:* *MAP3K7* variants are known to cause rare syndromic disease, including cardiospondylocarpofacial syndrome (CSCF). Some patients with CSCF develop dilated cardiomyopathy (DCM), however we do not know whether *MAP3K7* variants cause primary or isolated DCM without overt syndromic disease.

*Key Finding:* We present multiple orthogonal data sources supporting an association between *MAP3K7* loss-of-function variants and DCM. The collective evidence for this gene-disease association encompasses paediatric trio *de novo* discovery, GWAS, familial segregation, rare variant case-control analysis and variant categorisation *in vitro*. Strong segregation evidence was obtained for p.(Tyr125Cys), with nine clinically affected genotype-positive individuals in one pedigree (estimated LOD score 2.7). Further, p.(Cys266del) was observed in three affected individuals in one family and a *de novo* paediatric case.

*Take-Home Message:* Loss-of-function variants in *MAP3K7* are associated with DCM and several affected individuals lack overt syndromic features. *MAP3K7*-related disease may present as apparently isolated DCM across the lifespan. These findings support the incorporation of *MAP3K7* testing into DCM diagnostic pipelines. 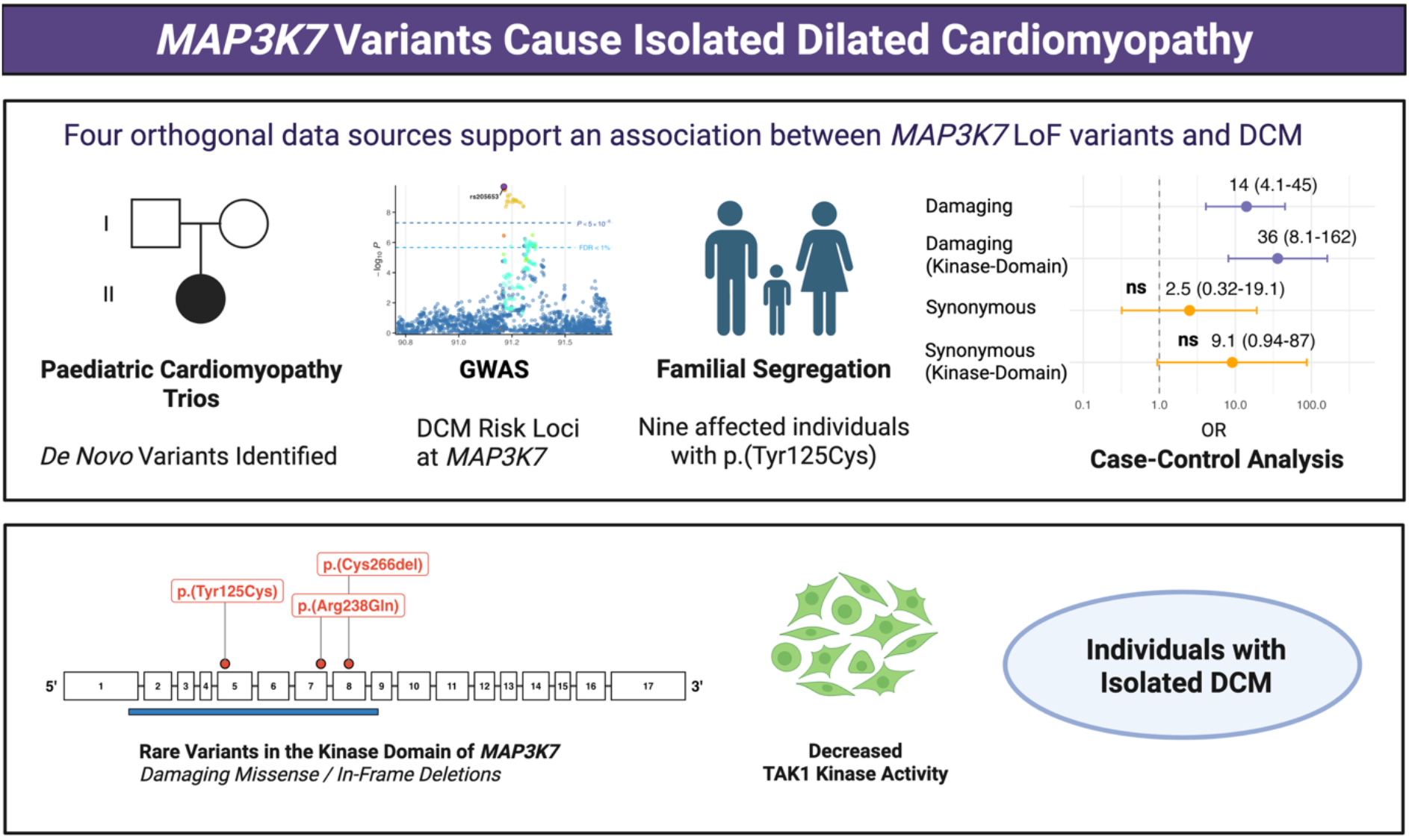

## INTRODUCTION

Dilated cardiomyopathy (DCM) is a chronic, often progressive, heritable disorder of the heart muscle, characterised by ventricular dilatation and systolic dysfunction not attributable to myocardial ischaemia or abnormal haemodynamic loading. It is a leading cause of heart failure and sudden cardiac death, and can present across the lifespan, with an estimated prevalence of approximately 1 in 250 adults^1^. In children it is much less common, with prevalence estimates of 0.026-0.046% depending on age (higher in infancy). DCM accounts for approximately 50% of all paediatric cardiomyopathy cases^2–5^. Genetic studies have demonstrated that DCM is highly heterogeneous, with pathogenic variants identified in a wide range of genes encoding structural, cytoskeletal, and regulatory proteins^6^. Despite these advances, many individuals with DCM remain without a genetic diagnosis, limiting opportunities for personalised clinical management and cascade genetic testing of at-risk relatives. In this study, we identify rare variants in *MAP3K7* as a novel cause of isolated DCM, and demonstrate loss-of-function as the underlying molecular mechanism.

*MAP3K7* (mitogen-activated protein kinase kinase kinase 7), encodes a serine/threonine kinase, known as TAK1 (Transforming growth factor-β-activated kinase 1). TAK1 is involved in multiple intracellular signalling pathways and is critical for cardiac cell survival and homeostasis. Activation of TAK1 involves binding to TAB1, TAB2, and TAB3, and regulates pathways including the JNK, MAPK-p38, ERK and NF-kB pathways^7–9^. Cardiac specific knockout of *MAP3K7* in mice induces cell death, pathological remodelling of the heart, and heart failure^10^.

In humans, variants in *MAP3K7* have been associated with two rare autosomal dominant syndromes, frontometaphyseal dysplasia type 2 (FMD2) due to gain of function missense variants,^11^ and cardiospondylocarpofacial syndrome (CSCF) associated with variant-specific loss of function (LoF)^7^. Notably, pathogenic variants associated with CSCF are predominantly missense or in-frame changes. Although a single frameshift variant has been reported^12^, the current evidence suggests that CSCF is primarily caused by altered TAK1 function rather than haploinsufficiency. Both FMD2 and CSCF include craniofacial, skeletal and congenital heart defects, highlighting the importance of TAK1 for skeletal and cardiac development^7,8,11^.

DCM has recently been described as part of the CSCF phenotype. However, to our knowledge, *MAP3K7-*related DCM has been reported exclusively in the context of syndromic disease^8,13–15^. Given that LoF variants in *MAP3K7* show variable expressivity and phenotypes differ even among carriers of the same pathogenic variant^8^, it is plausible that cardiac presentations may occur in the absence of overt syndromic features.

Here, we show that patients with *MAP3K7* LoF variants can present with a predominantly DCM phenotype, with other features of CSCF absent or sufficiently subtle to be overlooked. We present four converging lines of human genetic evidence implicating *MAP3K7* in DCM: (1) recurrent *de novo* variants in paediatric cardiomyopathy cases; (2) association of individual common variants with adult DCM susceptibility identified through GWAS^16,17^; (3) familial segregation; and (4) enrichment of rare damaging variants in DCM cases compared with controls. Finally, functional studies support a LoF mechanism for three rare disease-associated variants, consistent with DCM being part of the LoF-CSCF spectrum of *MAP3K7*-disease.

## METHODS

### Identification of de novo MAP3K7 variants in children with cardiomyopathy in the 100,000 Genomes Project cohort

Paediatric cardiomyopathy probands were identified from the 100,000 Genomes Project (100KGP) rare disease programme^18,19^. Inclusion criteria for this analysis comprised recruitment under a cardiomyopathy category or the presence of “cardiomyopathy” in any Human Phenotype Ontology (HPO) term. This cohort therefore included children with either isolated cardiomyopathy or those with cardiomyopathy as part of a broader phenotype. Childhood onset was defined as age ≤16 years at consent or diagnosis, age at onset between 1–16 years (excluding onset at 0 years unless meeting other criteria), or age ≤16 years at first cardiology appointment (hospital episode statistics (HES) outpatient codes 320/170). *De novo* variant (DNV) data were available for 13,949 trios in 100KGP. Of 134 potentially eligible paediatric cardiomyopathy probands with both parents available, 117 trios showed phenotype distribution potentially consistent with *de novo* inheritance (i.e., neither parent was reported as affected) and were included in the analysis.

DNVs were identified by Genomics England using the rare disease pipeline (v9), with alignment by Illumina iSAAC and joint variant calling using Platypus. Candidate DNVs were detected from Mendelian inconsistencies and annotated with Ensembl VEP. DNVs were extracted from the 100KGP LabKey table (*denovo_flagged_variants*, main programme v18, 2023-12-21), which incorporates corrections for problematic genomic regions. All analyses were conducted within the Genomics England Research Environment under approved access (REC 14/EE/1112).

DNV enrichment was assessed using the denovolyzeR package^20^, which models gene-specific mutation rates based on sequence context and gene size^21^, and compares observed versus expected counts using Poisson statistics. Enrichment of genes with multiple DNVs was assessed by randomly redistributing observed variants across genes according to mutability and comparing the number of genes with recurrent hits to an empirical distribution generated from 10,000 permutations. Gene-level enrichment of missense DNVs in cardiac expressed genes was evaluated using a Poisson test, with Bonferroni correction for multiple testing. Cardiac expressed genes were defined as the 4420 genes in the top quartile for expression in the developing mouse heart at embryonic day 14.5^22^. This gene set was chosen because genes contributing to early onset cardiomyopathy may be expressed in the developing heart. Further, these genes have previously shown enrichment for *de novo* variants in congenital heart disease trios^22,23^.

Additional individuals with variants of interest were identified through 100KGP, DECIPHER, and the published literature. Further phenotype information was obtained through clinical collaboration/re-contact where appropriate.

#### GWAS

A genome-wide association study meta-analysis of DCM was performed for 14,256 cases and 1,199,156 controls of European ancestry from 16 cohorts within the HERMES consortium, and has been previously reported^17^. Briefly, association testing included ∼9.6 million common variants (minor allele frequency >1%) using inverse-variance weighted fixed-effects meta-analysis. To increase power, multitrait analysis of GWAS (MTAG) incorporated genetically correlated cardiac magnetic resonance– derived traits from 36,203 UK Biobank participants. At each GWAS locus, likely effector genes were prioritised using integrative approaches including nearest gene, variant-to-gene mapping, transcriptome-wide association studies, and functional genomic annotation. Rare variant burden testing was carried out for 58 prioritised genes using exome and genome sequencing data from UK Biobank and Genomics England cohorts. Three genes, which included *MAP3K7,* not previously established as causes of cardiomyopathy showed significant associations.

### Go-DCM analysis

We analysed variants in *MAP3K7* using whole genome sequencing (WGS) data from 756 DCM patients who were part of the GO-DCM study. Adult patients with DCM were prospectively recruited from multiple centres across the UK. The study was approved by the NHS Health Research Authority (REC reference: 18/LO/0692). Sequencing reads were aligned to the masked hg38 reference genome using DRAGMAP (v1.2.1). Variant calling was performed using GATK HaplotypeCaller (v4.5.0) following best practices, with joint genotyping across samples. Variants were annotated using VEP (v111), ClinVar (2024-04-21), and gnomAD (v4.1.0). Candidate variants were restricted to rare damaging missense variants in the protein kinase domain of *MAP3K7* (NM_145331.3), defined as variants with MAF <0.0001 in gnomAD and REVEL score >0.5 affecting amino acid residues 36–291. The variant, c.374A>G, p.(Tyr125Cys), was identified in two related individuals with prior negative genetic testing.

### Family segregation

The two individuals with the c.374A>G, p.(Tyr125Cys) variant were contacted by their clinical teams and subsequently seven additional affected relatives were invited to the SMARTER CM study (REC reference: 22/ES/0044). Written informed consent was obtained, including consent for publication of clinical data. Clinical and imaging data were collected, and blood samples were obtained for DNA extraction. Targeted Sanger sequencing was performed using specific primers (forward sequence TTCAGGTCCCTGTGAATTAG; reverse sequence GTGCTTGCATTCACATTGTG). PCR amplification was carried out under standard conditions using Kod polymerase, and sequencing was performed by GENEWIZ. A previously confirmed carrier of the variant was included as a positive control, and an unrelated individual sequenced on the same platform was included as a negative control. Sequence traces were analysed to confirm variant presence and segregation within the family.

### Analysis of genomes from the Australian Genomics Cardiovascular Disorders Flagship Study

We analysed WGS data from 487 probands in the Australian Genomics Cardiovascular Disorders Flagship, a cohort of patients with inherited cardiomyopathies, arrhythmias and congenital heart diseases enrolled via cardiac genetic sites across Australia^24^. Raw sequencing data were aligned to GRCh38 and were analysed using the GATK best-practice joint-calling pipeline for single nucleotide variants (SNVs), indels, and structural variants (SV) with upload to the Centre of Population Genomics-instance of *seqr*, cloud-hosted in Australia for variant filtering and analysis^24,25^. Candidate variant analysis was restricted to rare variants in *MAP3K7*, defined as those present at <0.0001 MAF and in zero homozygotes in gnomAD v4, in probands with a diagnosis of DCM. We identified a rare in-frame single amino acid deletion: NM_145331.3*(MAP3K7)*: c.798_800del, p.(Cys266del) in a gene-elusive proband with peripartum DCM and an extensive family history of DCM. To rule out other potential causal variants, we reviewed all rare variants present in at least one allele in the proband’s genome across 555 genes with putative cardiac disease associations from PanelApp Australia. Rare SVs and SNVs with an Ensembl-VEP of high/moderate impact, or with a SpliceAI maximum raw delta score >0.10 were reviewed and excluded. The proband and family members were recruited to the Elusive Hearts study for further characterisation and variant segregation with WGS. Additional WGS data from affected siblings were processed and analysed as indicated above.

### HEK293T transfection

To assess the functional effects of the identified variants, *MAP3K7* wild-type and mutant constructs were generated in expression vectors (pcDNA3.1 from Genescript) and engineered with a C-terminal FLAG tag. Plasmids were sequence-verified prior to transient transfection into HEK293T cells. Cells were cultured in DMEM medium (Gibco #31966047) and supplemented with 10% FBS (Sigma-Aldrich), 100 U/mL penicillin and streptomycin. All cells were maintained at 37° and 5% CO2. Transient transfection was performed using polyethylenimine (PEI). The transfection rate was determined by control transfections using pmaxGFP (Adgene) and transfection efficiency determined to be 90-100% by imaging GFP expression by fluorescence microscopy.

Four independent transfection experiments were performed with the following constructs: *MAP3K7 ^WT^ ; MAP3K7 ^Cys266del^ ; MAP3K7 ^Tyr125Cys^ ; MAP3K7 ^Arg238Gln^ ; MAP3K7 ^Gly110Asp^* as a known loss of function control^8^; and *MAP3K7 ^Val100Glu^* as a known gain of function control^11^. Cells were lysed 48 hours post-transfection. Protein expression and downstream signalling were assessed by Western blotting. Cell lysate proteins were resolved by SDS-PAGE on 4-12% Bis-Tris gel (Invitrogen NuPAGE; NP0336) and transferred to PVDF membrane. Membranes were probed with the following primary antibodies: Phospho-TAK1 (Thr184) Cell Signalling #4537, Phospho-p44/42 MAPK (Erk1/2) (Thr202/Tyr204) Cell Signalling #9101, p44/42 MAPK (Erk1/2) Cell Signalling #4696, vinculin Merck#V9131. After extensive washing, membranes were incubated for 1 hour with secondary antibodies (IRDye 680RD goat anti-mouse IgG; IRDye 800CW goat anti-rabbit IgG; LI-COR Biosciences). Blots were imaged on a LI-COR Odyssey CLx.

### Case-control analysis for rare variant association

#### (i) Cases

*MAP3K7* DCM case-control analysis was performed. The DCM cohort comprised 1449 eligible participants from nine studies recruited from the UK, who all had detailed baseline phenotypic characterisation, and underwent harmonised genome sequencing: Go-DCM^26^ (NCT03843255), the Royal Brompton & Harefield Cardiovascular Research Centre Biobank^27^, MATCH and MATCH2^28^, REMIT DCM^29^ (NCT04957147), and the HeartHive Cardiomyopathy Study^30^ (NCT04612296) are all observational studies to understand the genetic architecture and natural history of heart failure (HF) and/or DCM that have been previously described. SMARTER-CM (NCT05750147) is an ongoing observational study of cardiomyopathies not previously reported with the first 30 participants with DCM included here. MitoDCM^31^ (NCT05410873) is an interventional study to evaluate MitoQ as an adjunct therapy in DCM, and TRED-HF^32^ (NCT02859311), and TRED-HF2 (NCT06091475) are interventional studies evaluating the safety of withdrawal of therapies in patients in remission from DCM. In all studies, DCM was defined as left ventricular (LV) dilatation with reduced systolic function in the absence of coronary disease or abnormal loading, as previously described^33^. Samples underwent PCR-free WGS using Illumina short read technology (Broad Institute Genomics Platform), with alignment and variant calling using either the Illumina DRAGEN platform or a custom-built DRAGEN-GATK equivalent pipeline.

#### (ii) Controls

Population control samples were obtained from the INTERVAL study^34^. WGS data were downloaded from the EGA archive and re-processed using the same DRAGEN-GATK variant calling pipeline applied for cases, and harmonised annotation. gnomAD v4.1 (a publicly accessible aggregated database of exome and genome sequencing data) was used as a population reference. Allele count, number and frequency information was obtained from this database for European genomes in gnomAD^35^. All participants in both the case and control groups gave written and informed consent.

#### (iii) Filtering and analysis

Variants in the INTERVAL and DCM cohorts were filtered to a DP (read depth) > 10 and a GQ (genotype quality) > 20. Variants in gnomAD were included if they passed the gnomAD quality controls filters for the genome and all variants were annotated using Ensembl Variant Effect Predictor (VEP) v114/115. Variants in *MAP3K7* were identified using the Matched Annotation from NCBI and EMBL-EBI (MANE) Select transcript NM_145331.3. Analyses were restricted to ultra-rare variants with a gnomAD v4.1.0 POPMAX allele frequency <1 × 10⁻⁵. Eligible variants for the non-truncating analysis included missense variants, in-frame insertions/deletions, and splice-region variants predicted to escape nonsense-mediated decay (NMD). Missense variants were further required to have a REVEL score >0.5. To capture carriers of variants in the protein kinase domain, variants were additionally filtered to amino acids 36–291. Synonymous variants were obtained as reported above with the exception of the consequence being synonymous and exclusion of the REVEL filtering step. Truncating variants were defined as variants annotated as frameshift, start-loss, stop-gained, splice acceptor, or splice donor variants, with frameshift and splice variants restricted to those predicted to trigger nonsense-mediated decay. Burden testing to assess the enrichment of variants in cases compared to controls was performed using the *fisher_exact()* function in Python’s *scipy.stats* package, comparing the number of variant carriers and non-carriers between cases and controls using a 2 × 2 contingency table.

### Graphical design

The figures for this publication were generated with the assistance of resources provided by Biorender and/or design in R and Inkscape.

## RESULTS

We present four complementary lines of human genetic evidence implicating *MAP3K7* as a novel cause of DCM: (1) enrichment of *de novo* variants in paediatric cardiomyopathy trios; (2) independent identification of *MAP3K7* as a novel DCM locus in adult GWAS; (3) segregation of rare variants with disease in two large families; and (4) enrichment of rare damaging variants in DCM cases compared with controls.

### 1. Identification of de novo MAP3K7 variants in the 100KGP Paediatric Cardiomyopathy Cohort

We investigated protein coding *de novo* variants in 117 paediatric cardiomyopathy trios in the 100KGP, a UK large-scale genomics study. These paediatric trios included probands with syndromic and isolated cardiomyopathy. We used a previously reported *de novo* expectation model to assess and compare variant rates by variant class^20,21^. We identified two probands with rare *de novo* missense variants in *MAP3K7*, significantly more than would be expected by chance (Bonferroni adjusted p = 0.036).

The first *de novo* variant, c.713G>A, p.(Arg238Gln) (*Family 1*) was identified in a proband presenting with DCM at <5 years of age without additional classical features of CSCF (**Table 1**). The second *de novo* variant, c.467A>T, p.(Asp156Val), was identified in a proband with paediatric-onset cardiomyopathy and additional syndromic features consistent with CSCF. Notably, the p.(Asp156Val) variant has been previously reported in an individual with CSCF and valvular heart disease without cardiomyopathy^36^. Both variants are absent from gnomAD (v4.1.0) and fall within the TAK1 protein kinase domain, which overlaps the TAB1 binding domain. This region is under marked missense constraint^35^ and all *MAP3K7* variants associated with CSCF fall within this region^8^.

**Table 1:** *MAP3K7* Variant Table. Variant and phenotypic details of patients with DCM carrying *MAP3K7* variants identified in this study, together with available clinical information from additional individuals carrying the same variants, either previously reported or subsequently identified.

| Variant | Variant Type, Location, Effect | Allele Frequency (gnomAD) | Case(s) | Inheritance | Cardiac Phenotype | Other Features |
| --- | --- | --- | --- | --- | --- | --- |
| **c.713G>A, p.(Arg238Gln)** | Missense, Kinase domain, LOF (*in vitro*) | Absent | Family 1 (100KGP) | *De novo* | Paediatric-onset DCM aged <5 years | Short stature |
|  |  |  | 100KGP Case 2 | *De novo* | Mitral valve prolapse with moderate mitral regurgitation, impaired RV function, borderline impaired LV function | Short stature, feeding difficulties, round-tipped nose, thickened supraorbital ridges, high arched palate, brachydactyly, joint laxity, sensorineural hearing loss, mild learning difficulties, testicular torsion |
|  |  |  | DECIPHER Case | *De novo* | Prenatal-onset DCM | Short stature, feeding difficulties, ptosis, widely spaced nipples, joint laxity, scoliosis, clinodactyly 5th finger, camptodactyly 5th finger, hypotonia, autism spectrum disorder with intellectual disability requiring specialist educational provision |
|  |  |  | Published Case<sup>8</sup> | *De novo* | Congenital heart defect and mitral valve prolapse | Short stature, feeding difficulties, ptosis, round-tipped nose and other facial features in keeping with CSCF, high arched palate, hypotonia, brachydactyly and conductive hearing loss |
| **c.374A>G, p.(Tyr125Cys)** | Missense, Kinase domain, LOF (*in vitro*) | Absent | Family 2 II:3 | Autosomal Dominant Inheritance | DCM diagnosed in 50s, ICD | Ptosis, epicanthal folds, anteverted nares |
|  |  |  | Family 2 II:4 |  | DCM diagnosed in 40s with myocardial fibrosis, ICD, LVEF 30-49%, LVEDD 5.6 | Hypertelorism |
|  |  |  | Family 2 III:3 |  | DCM diagnosed as an adult | Ptosis, epicanthic folds, periorbital fullness |
|  |  |  | Family 2 III:4 |  | DCM diagnosed in 20s with severe LV systolic dysfunction and NSVT, ICD, LVEDD 7.1, LVEF 26% | Full cheeks, periorbital fullness, anteverted nares, round-tipped nose |
|  |  |  | Family 2 III:5 |  | DCM diagnosed aged 15-20 years, LVEDD 5.3 | Low set ears, ptosis, periorbital fullness, anteverted nares, joint laxity |
|  |  |  | Family 2<br>III:6 |  | Mild DCM diagnosed in 30s,<br>LVEDD 4.3, LVEF 45% | Hypotonic face, low set ears, posteriorly rotated ears, periorbital fullness, round-tipped nose |
|  |  |  | Family 2<br>IV:1 |  | DCM diagnosed aged 15-20 years | Ptosis (mild), periorbital fullness, anteverted nares, round-tipped nose |
|  |  |  | Family 2<br>IV:2 |  | Paediatric-onset DCM diagnosed aged <10 years with biventricular dysfunction, RVEF 41%, non-dilated RV, mild-moderate LV dilatation | Full cheeks, low set ears, posteriorly rotated ears, periorbital fullness, anteverted nares, joint laxity |
|  |  |  | Family 2<br>IV:3 |  | Paediatric-onset DCM diagnosed aged 10-15 years, LVEDD 4.3, RVEF 41% | Hypotonic face, low set ears, posteriorly rotated ears, hypertelorism, ptosis, anteverted nares, joint laxity |
| **c.798_800del, p.(Cys266del)** | In-frame deletion, Kinase domain, LOF (*in vitro*) | Absent | Family 3:<br>II:1 | Autosomal Dominant Inheritance | DCM diagnosed in 20s. Deterioration of condition during pregnancy (LVEDD ~7cm, LVEF ~19%). | Mild brachydactyly |
|  |  |  | Family 3:<br>II:5 | Inherited | Mildly dilated LV and RV, LVEF 50%, RVEF 46% |  |
|  |  |  | Family 3:<br>II:8 | Inherited | Mildly dilated LV, LVEF 46% |  |
| **c.798_800del, p.(Cys266del)** | In-frame deletion, Kinase domain, LOF | Absent | Family 4 (ClinVar) | *De novo* | Paediatric-onset DCM | Seizures |
| **c.608-1G>A** | Splice acceptor variant<br>Effect unknown | Absent | Genomics England (Proband) | Inherited | DCM diagnosed in 30s | Not reported |
|  |  |  | Genomics England (Parent) | Unknown | DCM diagnosed in 60s | Not reported |
| **c.121-2A>G** | Splice acceptor variant<br>Effect unknown | Absent | Genomics England | Unknown | DCM diagnosed in 30s | Not reported |

**Abbreviations:** 100KGP, 100,000 Genomes Project; CSCF, cardiospondylocarpofacial syndrome; DCM, dilated cardiomyopathy; ICD, implantable cardioverter-defibrillator; LOF, loss of function; LV, left ventricle; LVEDD, left ventricular end-diastolic diameter; LVEF, left ventricular ejection fraction; NSVT, non-sustained ventricular tachycardia; RV, right ventricle; RVEF, right ventricular ejection fraction.

Further investigation revealed the first variant, p.(Arg238Gln), to be a recurrent *de novo* variant associated with variable phenotypic expression. In addition to the proband described here, the same variant has previously been reported in an individual with CSCF and congenital heart disease without cardiomyopathy^8^. We also identified the variant in an additional 100KGP participant who was not captured in the original analysis because cardiomyopathy was neither recorded as an HPO term nor listed as the recruitment indication. A further individual harbouring the variant has been reported in DECIPHER^37^ (**Table 1**). Together these individuals highlight the variable expressivity associated with *MAP3K7* variants.

### 2. GWAS

In parallel, but independent of the paediatric work above, two large genome-wide association studies of adult DCM, one of which was carried out by our group, identified a novel DCM risk locus implicating *MAP3K7*^16,17^ (**Figure 1**). Using an integrative gene prioritisation framework incorporating multiple complementary genomic predictors, *MAP3K7* was subsequently prioritised as the most likely effector gene at this locus^17^.

**Figure 1:**
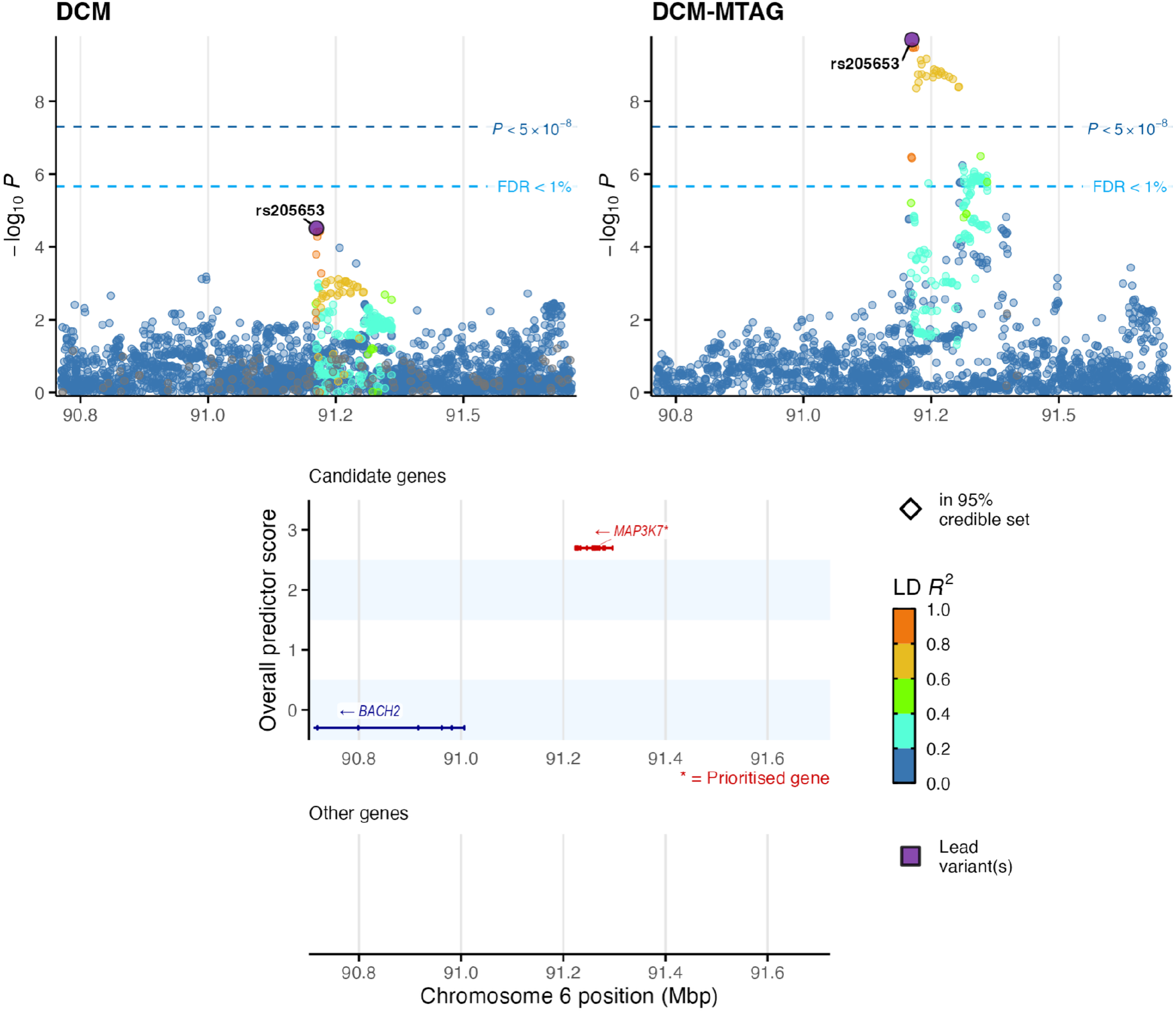
*MAP3K7* identified through DCM GWAS. Regional association plots of the chromosome 6 locus containing *MAP3K7* from the DCM GWAS^17^. Top panels: association statistics are shown for DCM and DCM-MTAG (multi-trait analysis combining DCM with cardiac magnetic resonance traits with high genetic correlation). Each point represents a genetic variant coloured according to linkage disequilibrium (LD; r²) with the lead variant, rs205653. Horizontal dashed lines indicate genome-wide significance (P < 5 × 10⁻⁸) and a false discovery rate (FDR) threshold of 1%. Bottom panel: Locus zoom plot showing *MAP3K7* as the sole overlapping gene at this position. In addition, the integrative gene-prioritisation framework previously prioritised *MAP3K7* as the likely effector gene at this locus^17^.

### 3. Familial segregation

The convergence of these independent rare and common variant findings on the same gene strongly supported *MAP3K7* as a candidate DCM gene and prompted us to examine two additional independent cohorts for further families harbouring rare *MAP3K7* variants. We identified two further DCM pedigrees harbouring rare *MAP3K7* variants (**Figure 2**, **Table 1**).

**Figure 2:**
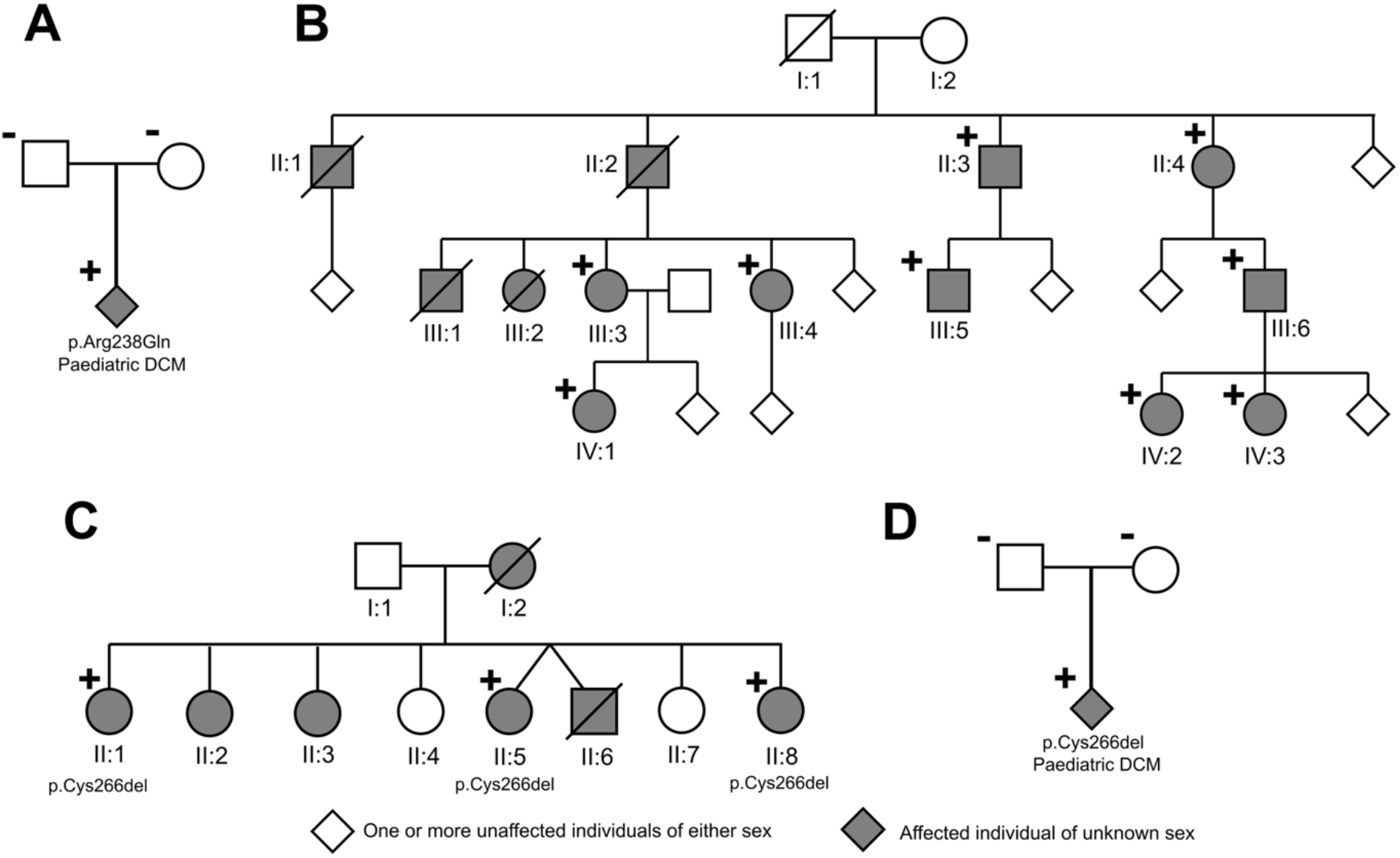
*MAP3K7* DCM Pedigrees. **A:** The p.Arg238Gln variant was identified in a *de novo* case of paediatric DCM. **B:** The p.Tyr125Cys variant was found in nine family members with a primary phenotype of DCM. On re-examination after identification of *MAP3K7* as a potential cause, subtle facial features consistent with CSCF were identified in all affected individuals and 3/9 also had joint laxity. **C:** The p.Cys266del variant was identified in three affected siblings in an Australian pedigree with an extensive history of DCM. On re-examination after identification of *MAP3K7,* the proband (II:1) showed no musculoskeletal, craniofacial or neurological features consistent with CSCF other than mild brachydactyly. **D:** The same p.Cys266del variant was reported in an unrelated *de novo* case of paediatric DCM with seizures from the NYCKidSeq study.

#### Family 2: DCM, c.374A>G, p.(Tyr125Cys)

In the UK GO-DCM cohort (n=756), we identified a rare missense variant, c.374A>G, p.(Tyr125Cys), in a mother and daughter with DCM. The variant is absent from gnomAD (v4.1.0) and is predicted to be damaging (REVEL=0.67). Subsequent cascade recruitment demonstrated segregation with DCM in an additional seven affected relatives, yielding nine genotype-positive individuals fulfilling diagnostic criteria for DCM across the pedigree (estimated LOD score 2.7). Age at diagnosis ranged from childhood to mid-adulthood. Although no family members had previously been recognised as syndromic, targeted clinical genetics assessment identified subtle craniofacial features in several affected individuals that overlapped with the CSCF spectrum (**Table 1**). These findings suggest that *MAP3K7*-associated disease may present predominantly as familial DCM with syndromic features that are sufficiently subtle to be missed without targeted assessment, or that the additional features may vary depending on age.

#### Family 3: DCM, c.798_800del, p.(Cys266del)

We reviewed WGS data from 487 probands in the Australian Genomics Cardiovascular Disorders Flagship study^24^. We identified a rare in-frame deletion, c.798_800del, p.(Cys266del), in a proband with peripartum DCM (*Family 3, II:1*). The variant is absent from gnomAD (v4.1.0) and is located within the *MAP3K7* kinase domain. No alternative genetic explanation for disease was identified (for further details see the **Supplementary Materials**). Following identification of the *MAP3K7* variant, the proband underwent targeted clinical genetics assessment which revealed mild brachydactyly. No other syndromic features strongly suggestive of CSCF syndrome were identified. The proband (II:1) has a strong family history of DCM, including a brother who was diagnosed in early adolescence, underwent cardiac transplantation in his early 20s and subsequently died from transplant rejection. Two genotype-positive sisters (II:5 and II:8) have also demonstrated mild ventricular dilatation and dysfunction on cardiac screening (**Figure 2C**).

#### Family 4: DCM, c.798_800del, p.(Cys266del)

Review of Family 3 led to identification of the same p.(Cys266del) variant, reported independently in ClinVar as a *de novo* variant in a child with DCM and seizures but no other syndromic features.

A summary of *MAP3K7* variants detected in families 1-4 together with 100KGP, DECIPHER and Genomics England are reported in **Table 1** below.

## 4. Rare variant burden analysis

### a. Non truncating variants

The families described above provide additional evidence supporting the role of rare *MAP3K7* variants in DCM. To evaluate whether this association extends beyond these individual families, we next performed a rare variant burden analysis in ancestry-matched DCM cases and controls.

We compared the frequency of ultra-rare (FAF <1×10⁻⁵) damaging non-truncating *MAP3K7* variants (in-frame indels and missense variants with REVEL >0.5) in European ancestry DCM cases (n=1449) and two European ancestry control cohorts comprising 3525 participants from the INTERVAL study and 39345 individuals from gnomAD.

Damaging non-truncating variants were significantly enriched in DCM cases compared with both control cohorts (OR 14–22), with stronger enrichment when restricted to the kinase domain (OR 22– 36), see **Table 2**. Notably, all qualifying damaging variants identified in cases were located within the *MAP3K7* kinase domain. In contrast, no enrichment of synonymous variants was observed, consistent with a selective enrichment of damaging *MAP3K7* variants in DCM cases.

**Table 2:** Enrichment of rare damaging non-truncating *MAP3K7* variants in DCM cases (n=1449) compared with ancestry-matched (INTERVAL n=3525) and population controls (gnomAD n=39345).

| Category | INTERVAL |  |  |  | gnomAD |  |  |
| --- | --- | --- | --- | --- | --- | --- | --- |
|  | Cases (+/-) | Controls (+/-) | OR (95% CI) | P value | Controls (+/-) | OR (95% CI) | P value |
| Damaging non-truncating |  |  |  |  |  |  |  |
| Gene-wide | 4/1445 | 0/3525 | 22*<br>(1.2–408) | $7.18 \times 10^{-3}$ | 8/39337 | 14<br>(4.1–45) | $6.24 \times 10^{-4}$ |
| Kinase domain | 4/1445 | 0/3525 | 22*<br>(1.2–408) | $7.18 \times 10^{-3}$ | 3/39342 | 36<br>(8.1–162) | $5.09 \times 10^{-5}$ |
| Synonymous |  |  |  |  |  |  |  |
| Gene-wide | 1/1448 | 4/3521 | 0.61<br>(0.07–5.4) | 1.00 | 11/39334 | 2.5<br>(0.32–19.1) | 0.35 |
| Kinase domain | 1/1448 | 1/3524 | 2.43<br>(0.15–39) | 0.50 | 3/39342 | 9.1<br>(0.94–87) | 0.14 |

Damaging non-truncating variants were defined as missense variants with REVEL >0.5 or in-frame indels, both with filtering allele frequency (FAF) <1×10⁻⁵. Cases (+/-): number of cases with (+) and without (-) a variant; Controls (+/-): number of controls with (+) and without (-) a variant. OR, odds ratio; CI, confidence interval. P values were calculated using Fisher’s exact test. *Haldane–Anscombe corrected odds ratio

### b. Truncating variants

In our previous GWAS analyses^17^, we performed a rare variant burden analysis of protein-truncating variants (PTVs) in *MAP3K7* using Genomics England and UK Biobank data. We identified an enrichment of PTVs in DCM cases (odds ratio 24.2, adjusted P = 0.02). The associated variants comprised two canonical splice-site variants (c.608-1G>A and c.121-2A>G), one of which was identified in two related individuals. Carriers of truncating *MAP3K7* variants in the UK Biobank also demonstrated adverse cardiac remodelling, including increased left ventricular end-diastolic volume (+54 ml, adjusted P = 0.01) and end-systolic volume (+38 ml, adjusted P = 4.4 × 10⁻⁴). We also investigated the prevalence of truncating *MAP3K7* variants in our DCM cohort (n =1449), but identified no qualifying variants. Similarly, truncating variants were absent in the INTERVAL controls (0/3525) and were extremely rare in gnomAD controls (2/39345), limiting our ability to independently assess the previously reported association.

### Functional Categorisation of MAP3K7 Variants

To assess the effect of the three *MAP3K7* variants identified in our families, we transfected these variants into HEK293T cells and used western blotting to measure autophosphorylated TAK1 (pTAK1) and phosphorylated ERK1/2 (pERK1/2) as a downstream marker of TAK1 kinase activity. Vinculin was used as a loading control. All patient variants analysed: p.Cys266del, p.Tyr125Cys, p.Arg238Gln, and the previously reported CSCF variant, p.Gly110Asp, resulted in undetectable levels of pTAK1 and pERK1/2, indicative of reduced kinase activity. In contrast, cells transfected with wild-type protein exhibited detectable levels of pTAK1 and pERK1/2, and cells transfected with the gain-of-function p.Val100Glu variant displayed a pronounced increase in pTAK1 and pERK1/2 relative to the wild-type control (**Figure 3)**. These findings suggest that the variants identified in families with DCM demonstrate a LoF mechanism like that reported for other CSCF-associated variants. This is consistent with the observation that some patients with CSCF develop DCM (**Figure 4, Supplementary Table 3**)^8^ whereas there are no reports of DCM in patients with FMD2.

**Figure 3:**
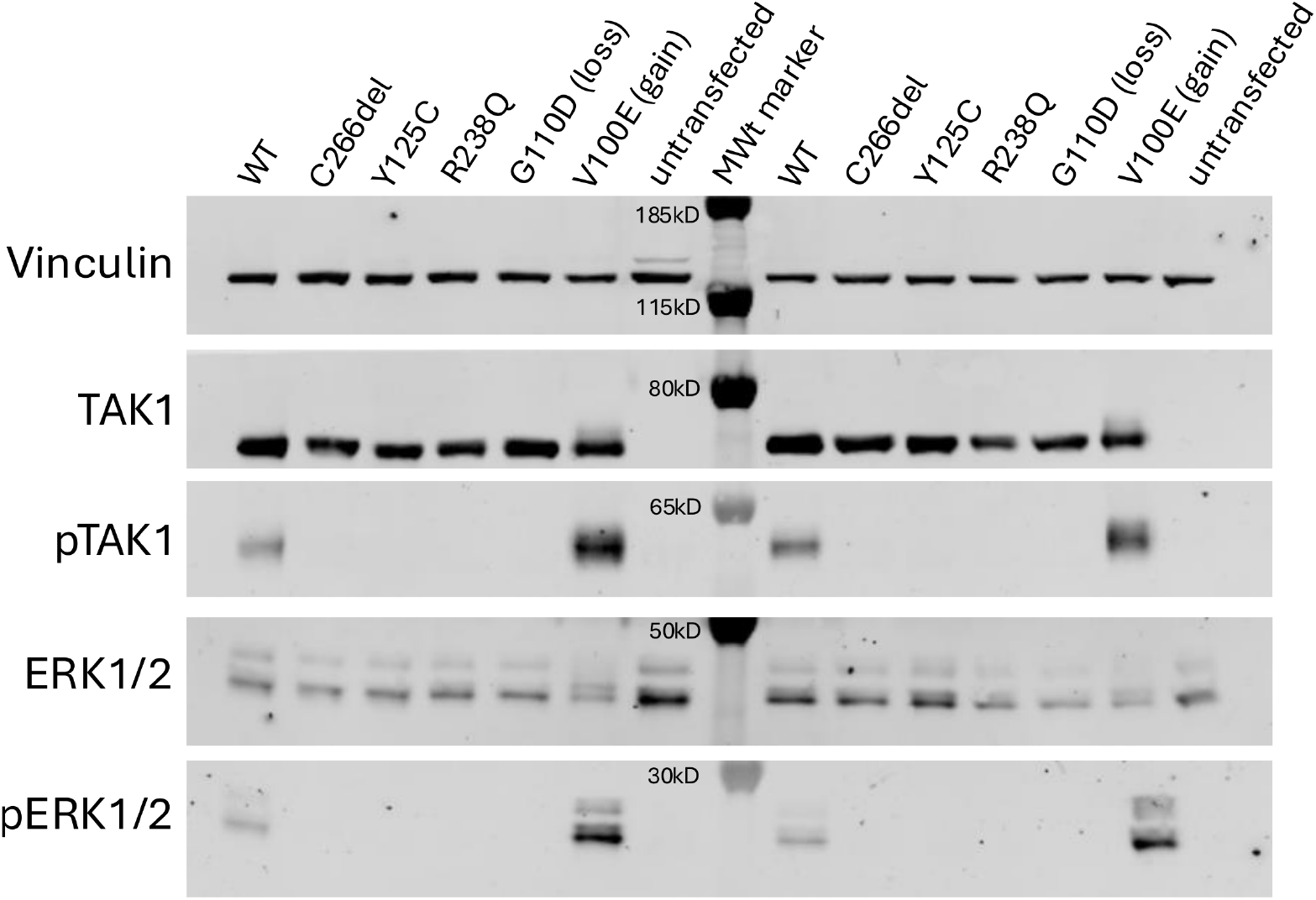
Evaluation of *MAP3K7* Variants. Western blots from two representative experiments (out of four). Rare DCM associated *MAP3K7* variants result in decreased TAK1 kinase activity, with decreased autophosphorylation at TAK1 residue Thr184 and decreased ERK1/2 phosphorylation.

**Figure 4:**
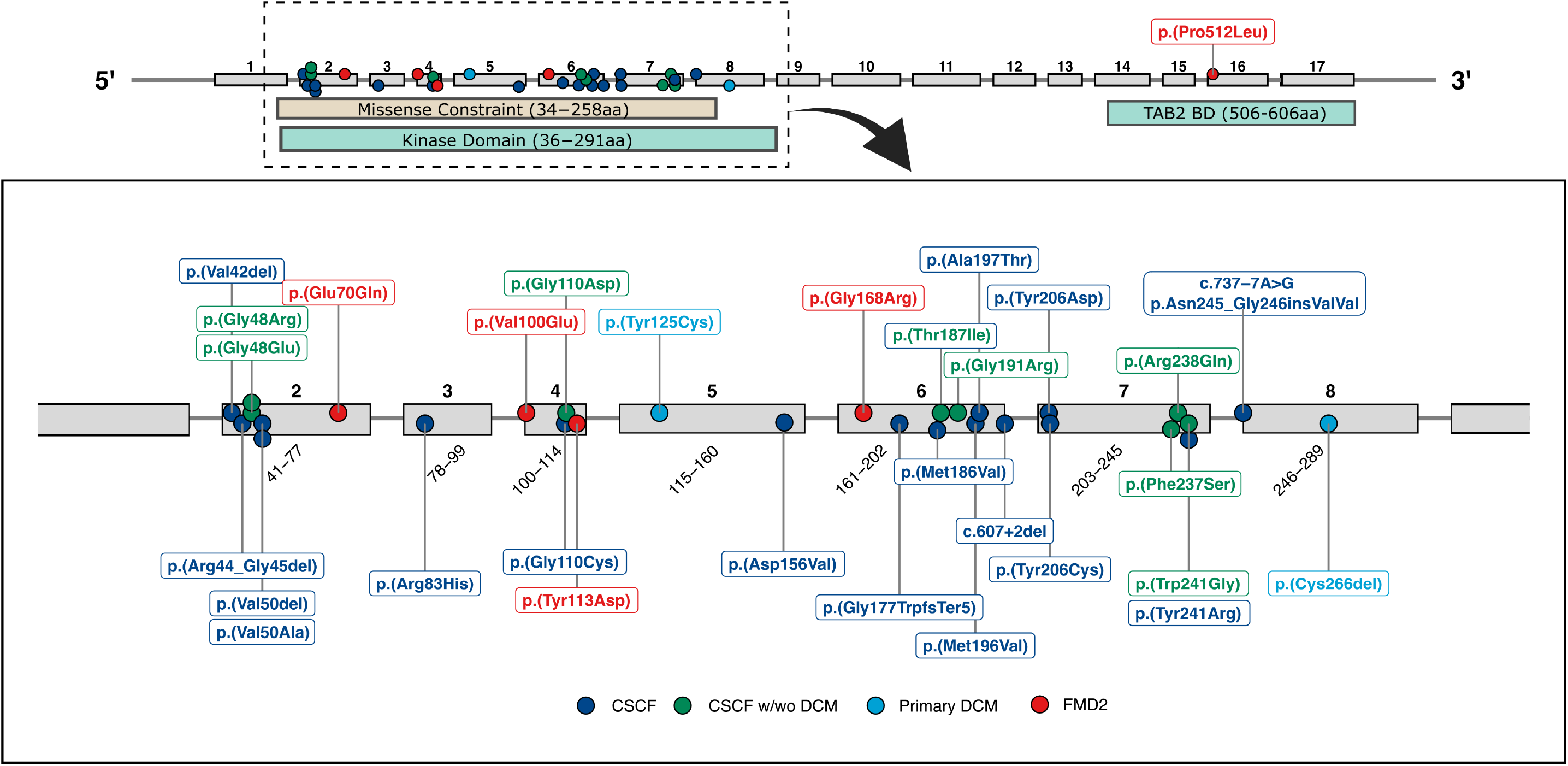
*MAP3K7* Variants Associated with CSCF, DCM or FMD2. Heterozygous *MAP3K7* variants reported in patients with cardiospondylocarpofacial syndrome (CSCF), primary dilated cardiomyopathy (DCM) or frontometaphseal dysplasia type 2 (FMD2) depicted in the MANE Select transcript NM_145331.3. We detected the p.Tyr125Cys and p.Cys266del variants in families with a primary DCM phenotype (light blue). The p.Arg238Gln, p.Gly48Glu, p.Gly110Asp, and p.Trp241Arg variants have been reported in patients with CSCF, with DCM documented in at least one patient harbouring each variant (green). The p.Thr187Ile variant (green) was reported in a patient with CSCF and paediatric heart failure who died from an arrhythmic event^48^.

## DISCUSSION

CSCF, a developmental disorder characterised by craniofacial, skeletal and cardiac abnormalities, is predominantly caused by missense or in-frame insertions/deletion variants within the kinase domain of *MAP3K7* that result in loss of function^8,38^. While DCM is a recently described feature of CSCF^8,13,14^, there have been no previous reports of isolated DCM attributed to *MAP3K7*. Here, we show that *MAP3K7* variation can present as primary DCM in the absence of overt syndromic features. To demonstrate this, we combine evidence from four independent lines of human genetic evidence: a *de novo* variant analysis, GWAS, familial segregation data, and rare variant burden testing. Our findings expand the phenotypic spectrum of *MAP3K7*-related disorders beyond classical CSCF and suggest that DCM may represent the predominant or sole manifestation of disease in some individuals. Together, these data support *MAP3K7* as a rare cause of apparently monogenic DCM in both paediatric and adult patients.

Critically, transfection studies in HEK293T cells demonstrated that the patient DCM variants, p.Cys266del, p.Tyr125Cys, and p.Arg238Gln, decrease TAK1 kinase activity *in vitro*. This is consistent with the LoF mechanism previously reported for other CSCF-associated variants^8,38^. Notably, one recurrent variant, p.(Arg238Gln), is associated with a variable phenotype, including both isolated DCM, and CSCF without cardiomyopathy^8^. This phenotypic overlap suggests that there may be no clear mechanistic distinction between variants associated with CSCF and those causing isolated DCM. However, larger cohorts will be required to examine this theory in more detail and identify any potential modifiers of disease expression.

A growing number of pathogenic variants in *MAP3K7* have been reported in patients with CSCF or FMD2 (**Figure 4**). Notably, all reported CSCF-associated variants are situated within the kinase domain. Four FMD2-associated variants are also located within the kinase domain, whereas one recurrent variant, p.(Pro512Leu), associated with a more severe FMD2 phenotype, lies within the TAB2-binding domain^11^. Reported pathogenic *MAP3K7* variants predominantly comprise missense variants, in-frame deletions and splice-altering variants. Although a single frameshift variant has been reported in an individual with CSCF and overlapping features of other MAP3K7-related disorders^12^, no functional studies have confirmed that the variant results in nonsense mediated decay (NMD) or haploinsufficiency. In contrast, a splice variant reported by Morlino et al. was shown to create a novel splice acceptor site, resulting in an in-frame insertion at the protein level^38,39^. Overall, the available evidence suggests that *MAP3K7*-associated CSCF is primarily driven by variant-specific decrease in TAK1 function rather than reduced gene dosage.

The PTV burden signal we previously identified was driven by two canonical splice-site variants (c.608-1G>A and c.121-2A>G)^17^. The transcript-level consequences of these variants have not yet been determined, and it therefore remains unclear whether they result in loss of function through NMD, altered splicing with preservation of the reading frame, or another mechanism. Further transcript and functional studies will be required to clarify how these variants contribute to disease.

At the molecular level, *MAP3K7* encodes the kinase TAK1, which functions in a signalling complex with TAB proteins (TAB1-3) to regulate downstream stress and inflammatory pathways^40,41^. Notably, *TAB2* LoF variants cause a syndrome reminiscent of CSCF, characterised by mitral valve disease, short stature, DCM and connective tissue disease^42–45^. Additionally, in mice, cardiomyocyte specific deletion of *Tab2* causes DCM with increased necrosis and apoptosis^45^. We suggest that disrupted TAK1-TAB2 signalling, due to LoF variants in either *MAP3K7* or *TAB2,* drives a shared pathology that may predispose to the development of cardiomyopathy. One proposed mechanism involves aberrant RIPK1 activation due to decreased TAK1 phosphorylation, driving cardiomyocyte death through pro-apoptotic and necroptotic pathways^45^. However, the precise molecular basis of cardiac involvement, and the factors determining variable expressivity, remain incompletely understood and warrant further investigation.

The findings from this work have important implications for clinical genetic testing. At present, *MAP3K7* is included on some syndromic panels, such as the R135 paediatric or syndromic cardiomyopathy panel in the UK^46^, but it is not consistently represented on standard DCM gene panels. We suggest that *MAP3K7* variants can present as apparently isolated DCM in both paediatric and adult populations, with extra-cardiac manifestations that may be absent, subtle, or only recognised retrospectively. This is exemplified by the large family with DCM identified in this study (Family 2, Figure 2B), which remained genetically unexplained for more than 20 years despite long-term clinical follow-up and assessment by clinical genetics. These observations suggest that *MAP3K7-*associated DCM may be under-recognised in routine diagnostic practice. Inclusion of *MAP3K7* in DCM-focused gene panels could therefore improve diagnostic yield and facilitate more accurate genetic counselling.

More broadly, our findings add to a growing recognition that the phenotypic boundaries between syndromic and non-syndromic cardiovascular disease can be blurred. Similar to observations in the RASopathy genes, where pathogenic variants may result in a spectrum of disease ranging from classical syndromic presentations to predominantly cardiac phenotypes^47^, we suggest that *MAP3K7*-associated disease also exists along a clinical spectrum ranging from classical CSCF to presentations characterised predominantly by DCM. This raises the possibility that additional genes regarded primarily as causes of developmental or syndromic disorders may also contribute to cardiomyopathy, particularly when extracardiac features are mild, overlooked, or incompletely penetrant. Larger studies in diverse cohorts will be important to define the contribution of *MAP3K7* variants to DCM more precisely.

## CONCLUSION

The findings presented here suggest that non-truncating *MAP3K7* variants associated with reduced kinase activity may represent a rare cause of autosomal dominant DCM, and affected patients may present without overt syndromic features. While some patients in our cohort exhibited subtle features consistent with CSCF, their primary presentation was cardiac. These observations expand the clinical spectrum of *MAP3K7*-related disorders to include apparently isolated DCM. This has important implications for genetic testing strategies and highlights a role for TAK1 signalling in cardiac disease.

To assess the effect of the three *MAP3K7* variants identified in our families, we transfected these variants into HEK293T cells and used western blotting to measure autophosphorylated TAK1 (pTAK1) and phosphorylated ERK1/2 (pERK1/2) as a downstream marker of TAK1 kinase activity. Vinculin was used as a loading control. All patient variants analysed: p.Cys266del, p.Tyr125Cys, p.Arg238Gln, and the previously reported CSCF variant, p.Gly110Asp, resulted in undetectable levels of pTAK1 and pERK1/2, indicative of reduced kinase activity. In contrast, cells transfected with wild-type protein exhibited detectable levels of pTAK1 and pERK1/2, and cells transfected with the gain-of-function p.Val100Glu variant displayed a pronounced increase in pTAK1 and pERK1/2 relative to the wild-type control (**Figure 3)**. These findings suggest that the variants identified in families with DCM demonstrate a LoF mechanism like that reported for other CSCF-associated variants. This is consistent with the observation that some patients with CSCF develop DCM (**Figure 4, Supplementary Table 3**)^8^ whereas there are no reports of DCM in patients with FMD2.

## Data Availability

Data from the National Genomic Research Library (NGRL) used in this research are available within the secure Genomics England Research Environment. Access to NGRL data is restricted to adhere to consent requirements and protect participant privacy. Data used in this research include: rare disease programme data from the 100,000 Genomes Project within the NGRL, including participant phenotype and clinical data used to identify paediatric cardiomyopathy probands and family relationships used to identify eligible parent offspring trios. De novo variant data were obtained from the Genomics England rare disease pipeline (v9), using the LabKey denovo flagged variants table (main programme v18, 2023-12-21). Derived datasets generated for this study comprised the eligible paediatric cardiomyopathy cohort and de novo variant datasets used for gene-level and enrichment analyses. These derived datasets remain within the secure Genomics England Research Environment. File paths: re_gecip/cardiovascular/imperial_paed_CM/Kat/denovo_analysis/VEP111/sample_ids; re_gecip/cardiovascular/imperial_paed_CM/Kat/denovo_analysis/VEP111/dnv_data. Access to NGRL data is provided to approved researchers who are members of the Genomics England Research Network, subject to institutional access agreements and research project approval under participant-led governance. For more information on data access, visit: https://www.genomicsengland.co.uk/research. Other data produced in the present study are available upon reasonable request to the authors and following appropriate ethics and legal approval.

## ACKNOWLEDGEMENTS

We sincerely thank the families who participated in this research for their significant contributions to advancing the scientific understanding of rare and inherited genetic diseases.

We gratefully acknowledge the participants of the National Genomic Research Library (NGRL), whose contributions made this research possible. Secure access to the NGRL under project ID 125 was provided by Genomics England, which delivers the NGRL in partnership with NHS England, and is wholly owned by the UK Department of Health and Social Care. The NGRL contains participants’ health data collected by the NHS as part of their care, along with samples and data from their participation in research, for which fully informed consent has been obtained. This includes genomic and clinical data provided through the NHS Genomic Medicine Service, as well as data obtained through research studies, including the 100,000 Genomes Project and the Generation Study, both of which are delivered in partnership with the NHS, and from other research cohorts involving external collaborators.

This research has been conducted using the UK Biobank Resource under Application Number 47602.

Participants in the INTERVAL randomised controlled trial were recruited with the active collaboration of NHS Blood and Transplant England (www.nhsbt.nhs.uk), which has supported field work and other elements of the trial. DNA extraction and genotyping was co-funded by the National Institute for Health Research (NIHR), the NIHR BioResource (http://bioresource.nihr.ac.uk/) and the NIHR [Cambridge Biomedical Research Centre at the Cambridge University Hospitals NHS Foundation Trust].* Sequencing data was supported by the Wellcome Trust grant number 206194. The academic coordinating centre for INTERVAL was supported by core funding from: NIHR Blood and Transplant Research Unit in Donor Health and Genomics (NIHR BTRU 201410024), UK Medical Research Council (MR/L003120/1), British Heart Foundation (RG/13/13/30194) and the NIHR [Cambridge Biomedical Research Centre at the Cambridge University Hospitals NHS Foundation Trust]. A complete list of the investigators and contributors to the INTERVAL trial is provided^34^. The academic coordinating centre would like to thank blood donor centre staff and blood donors for participating in the INTERVAL trial.

*The views expressed are those of the authors and not necessarily those of the NHS, the NIHR or the Department of Health and Social Care

## ETHICAL APPROVALS

All studies contributing to the DCM case–control cohort were conducted with appropriate ethical approval and participant consent, as detailed in the original study publications referenced in the Methods.

Relevant ethical approvals for the additional individual cases were obtained under the relevant study protocols. The GO-DCM study was approved by the NHS Health Research Authority (REC reference: 18/LO/0692). HRA and Health and Care Research Wales (HCRW) approval was obtained for the ICONIK study (REC reference: 22/NS/0025) and SMARTER CM study (REC reference: 22/ES/0044). Written informed consent was obtained for Family 2, including consent for publication of clinical data. Individual-level clinical information was reported in accordance with the consent provisions of the relevant study protocols; where additional phenotypic information was required, participants were recontacted and appropriate additional consent for reporting this information was obtained. Written informed consent for the Elusive Hearts study, approved by Royal Children’s Hospital Human Research Ethics Committee (97521) was obtained for four family members from Family 3. The consent included the publication of clinical phenotypic data.

## FUNDING

Analysis was supported by the Centre for Population Genomics (Garvan Institute of Medical Research and Murdoch Children’s Research Institute) and was funded in part by a National Health and Medical Research Council (NHMRC) investigator grant (2009982) and the Medical Research Future Fund (MRFF) Genomics Health Futures Mission (2032931). This work also forms part of Australian BioCommons’ GUARDIANS program, which is enabled by NCRIS investment via Bioplatforms Australia. The Australian Genomics Cardiovascular Genetic Disorders Flagship was funded by the Medical Research Future Fund (EPCD000028). Australian Genomics was funded by The National Health and Medical Research Council (NHMRC; GNTIER 1113531 and GNTIER 2000001). The Elusive Hearts study is funded by MRFF Cardiovascular Health Mission (2024269).

This work was supported by: Sir Jules Thorn Charitable Trust [21JTA], Wellcome Trust [222883/Z/21/Z]; [222883/Z/21/Z]; Medical Research Council (UK) [MC_UP_1605/13], British Heart Foundation [SP/17/11/32885; FS/CRLF/21/23011; BBC/F/21/220106; RE/24/130023; FS/IPBSRF/22/27059; RG/F/24/110138; CH/F/24/90015; FS/ICRF/24/26128], Rosetrees Trust, Pathfinder Cardiogenomics programme of the European Innovation Council of the European Union [DCM-NEXT: 101115416], Royal Brompton & Harefield Hospitals Charity, and the NIHR Imperial Biomedical Research Centre.

## DISCLOSURE OF INTEREST

JSW has received research support from Bristol Myers Squibb, has acted as a paid advisor to Health Lumen, Tenaya Therapeutics, Solid Biosciences, Hopkins Van Mil on behalf of Genomics England, and HEOR Ltd on behalf of Rocket Pharmaceuticals, and is a founder with equity in Saturnus Bio. JI is a consultant for Kardigan. BPH has received fees for consulting and advisory work from Astra Zeneca, Eli Lilly, Isomab, Novo Nordisk and Zoll.

## DATA AVAILABILITY STATEMENT

Data from the National Genomic Research Library (NGRL) used in this research are available within the secure Genomics England Research Environment. Access to NGRL data is restricted to adhere to consent requirements and protect participant privacy. Data used in this research include: rare disease programme data from the 100,000 Genomes Project within the NGRL, including participant phenotype and clinical data used to identify paediatric cardiomyopathy probands and family relationships used to identify eligible parent–offspring trios. *De novo* variant data were obtained from the Genomics England rare disease pipeline (v9), using the LabKey denovo_flagged_variants table (main programme v18, 2023-12-21). Derived datasets generated for this study comprised the eligible paediatric cardiomyopathy cohort and *de novo* variant datasets used for gene-level and enrichment analyses. These derived datasets remain within the secure Genomics England Research Environment. File paths: re_gecip/cardiovascular/imperial_paed_CM/Kat/denovo_analysis/VEP111/sample_ids; re_gecip/cardiovascular/imperial_paed_CM/Kat/denovo_analysis/VEP111/dnv_data. Access to NGRL data is provided to approved researchers who are members of the Genomics England Research Network, subject to institutional access agreements and research project approval under participant-led governance. For more information on data access, visit: https://www.genomicsengland.co.uk/research.

For information on data and code availability from the GWAS see Zheng et al.^17^. Other data produced in the present study are available upon reasonable request to the authors and following appropriate ethics and legal approval.

For the purpose of open access, the authors have applied a Creative Commons Attribution (CC BY) licence to any Author Accepted Manuscript version arising.

## Notes

### Competing Interest Statement

JSW has received research support from Bristol Myers Squibb, has acted as a paid advisor to MyoKardia, Pfizer, Foresite Labs, Health Lumen, Tenaya Therapeutics, Solid Biosciences, and Genomics England, and is a founder with equity in Saturnus Bio. JI is a consultant for Kardigan. BPH has received fees for consulting and advisory work from Astra Zeneca, Eli Lilly, Isomab, Novo Nordisk and Zoll.

### Author Declarations

The Human Research Ethics Committee of the Royal Children's Hospital gave ethical approval for the Elusive Hearts study (97521). The NHS Health Research Authority gave ethical approval for the GO-DCM study (REC reference: 18/LO/0692). HRA and Health and Care Research Wales (HCRW) approval was obtained for the ICONIK study (REC reference: 22/NS/0025) and the SMARTER CM study (REC reference: 22/ES/0044). This research has been conducted using the UK Biobank Resource under Application Number 47602. Secure access to the National Genomic Research Library (NGRL) under project ID 125 was provided by Genomics England, which delivers the NGRL in partnership with NHS England, and is wholly owned by the UK Department of Health and Social Care. The NGRL contains participants' health data collected by the NHS as part of their care, along with samples and data from their participation in research, for which fully informed consent has been obtained. This includes genomic and clinical data provided through the NHS Genomic Medicine Service, as well as data obtained through research studies, including the 100,000 Genomes Project and the Generation Study, both of which are delivered in partnership with the NHS, and from other research cohorts involving external collaborators.

### Summary of Updates

This version of the manuscript has been revised to incorporate updated clinical and genomic data from one of the families as well as an updated case-control analysis.

## REFERENCES

1. McGurk, K. A. et al. The penetrance of rare variants in cardiomyopathy-associated genes: A cross-sectional approach to estimating penetrance for secondary findings. The American Journal of Human Genetics 110, 1482–1495 (2023).

2. Lipshultz, S. E. et al. The Incidence of Pediatric Cardiomyopathy in Two Regions of the United States. New England Journal of Medicine 348, 1647–1655 (2003).

3. Nugent, A. W. et al. The Epidemiology of Childhood Cardiomyopathy in Australia. New England Journal of Medicine 348, 1639–1646 (2003).

4. Arola, A. et al. Epidemiology of Idiopathic Cardiomyopathies in Children and Adolescents: A Nationwide Study in Finland. Am. J. Epidemiol. 146, 385–393 (1997).

5. Andrews, R. E., Fenton, M. J., Ridout, D. A. & Burch, M. New-Onset Heart Failure Due to Heart Muscle Disease in Childhood. Circulation 117, 79–84 (2008).

6. Jordan, E. et al. Evidence-Based Assessment of Genes in Dilated Cardiomyopathy. Circulation 144, 7–19 (2021).

7. Le Goff, C. et al. Heterozygous Mutations in MAP3K7, Encoding TGF-β-Activated Kinase 1, Cause Cardiospondylocarpofacial Syndrome. The American Journal of Human Genetics 99, 407–413 (2016).

8. van Woerden, G. M. et al. The MAP3K7 gene: Further delineation of clinical characteristics and genotype/phenotype correlations. Hum. Mutat. 43, 1377–1395 (2022).

9. Xu, Y.-R. & Lei, C.-Q. TAK1-TABs Complex: A Central Signalosome in Inflammatory Responses. Front. Immunol. 11, 608976 (2021).

10. Li, L. et al. Transforming growth factor βactivated kinase 1 signaling pathway critically regulates myocardial survival and remodeling. Circulation 130, 2162–2172 (2014).

11. Wade, E. M. et al. Mutations in MAP3K7 that Alter the Activity of the TAK1 Signaling Complex Cause Frontometaphyseal Dysplasia. The American Journal of Human Genetics 99, 392–406 (2016).

12. Shepherd, W. B., Colaiacovo, S., Campbell, C. & Saleh, M. A novel MAP3K7 mutation in a child with cardiospondylocarpofacial syndrome and orofacial clefting. Clin. Genet. 103, 254– 255 (2023).

13. Das, B. B. & Criscuolo, J. J. Neonatal dilated cardiomyopathy and cardiospondylocarpofacial syndrome linked to a novel MAP3K7 gene mutation. Ann. Pediatr. Cardiol. 18, 68–71 (2025).

14. Yang, Q. et al. Genetic diagnosis and clinical characteristics analysis of cardiospondylocarpofacial syndrome in a Chinese family. Front. Pediatr. 13, (2025).

15. Schuermans, N. et al. Shortcutting the diagnostic odyssey: the multidisciplinary Program for Undiagnosed Rare Diseases in adults (UD-PrOZA). Orphanet J. Rare Dis. 17, (2022).

16. Jurgens, S. J. et al. Genome-wide association study reveals mechanisms underlying dilated cardiomyopathy and myocardial resilience. Nat. Genet. 56, 2636–2645 (2024).

17. Zheng, S. L. et al. Genome-wide association analysis provides insights into the molecular etiology of dilated cardiomyopathy. Nat. Genet. 56, 2646–2658 (2024).

18. Josephs, K. S. et al. Cardiomyopathies in 100,000 genomes project: interval evaluation improves diagnostic yield and informs strategies for ongoing gene discovery. Genome Med. 16, 125 (2024).

19. Genomics England. National Genomic Research Library Governance Framework. Figshare (2025).

20. Ware, J. S., Samocha, K. E., Homsy, J. & Daly, M. J. Interpreting *de novo* Variation in Human Disease Using denovolyzeR. Curr. Protoc. Hum. Genet. 87, (2015).

21. Samocha, K. E. et al. A framework for the interpretation of de novo mutation in human disease. Nat. Genet. 46, 944–950 (2014).

22. Homsy, J. et al. De novo mutations in congenital heart disease with neurodevelopmental and other congenital anomalies. Science (1979). 350, 1262–1266 (2015).

23. Zaidi, S. et al. De novo mutations in histone-modifying genes in congenital heart disease. Nature 498, 220–223 (2013).

24. Austin, R. et al. A multitiered analysis platform for genome sequencing: Design and initial findings of the Australian Genomics Cardiovascular Disorders Flagship. Genetics in Medicine Open 2, 101842 (2024).

25. Welland, M. J., et al. Scalable automated reanalysis of genomic data in research and clinical rare disease cohorts. medRxiv 7, (2025).

26. Mangino, M. et al. Sex Differences in the Genetic Causes of Dilated Cardiomyopathy. J. Am. Coll. Cardiol. 86, 400–403 (2025).

27. Mazzarotto, F. et al. Reevaluating the Genetic Contribution of Monogenic Dilated Cardiomyopathy. Circulation 141, 387–398 (2020).

28. Helbitz, A. et al. Identification, Characterisation and Outcomes of Pre-Atrial Fibrillation in Heart Failure with Reduced Ejection Fraction. ESC Heart Fail. 12, 3688–3696 (2025).

29. Hammersley, D. J. et al. Residual structural, functional and myocardial tissue abnormalities in patients with dilated cardiomyopathy in clinical remission. in Abstracts A18–A19 (BMJ Publishing Group Ltd and British Cardiovascular Society, 2024).

30. Roberts, A. M. et al. The Heart Hive: direct-to-participant recruitment provides an effective, scalable route to identify engaged participants for cardiomyopathy research. medRxiv (2025).

31. Halliday, B. P. et al. A double-blind, randomized placebo-controlled trial examining the effect of MitoQ on myocardial energetics in patients with dilated cardiomyopathy. Eur. Heart J. Cardiovasc. Imaging 27, 74–77 (2026).

32. Halliday, B. P. et al. Withdrawal of pharmacological treatment for heart failure in patients with recovered dilated cardiomyopathy (TRED-HF): an open-label, pilot, randomised trial. The Lancet 393, 61–73 (2019).

33. Arbelo, E. et al. 2023 ESC Guidelines for the management of cardiomyopathies. Eur. Heart J. 44, 3503–3626 (2023).

34. Di Angelantonio, E. et al. Efficiency and safety of varying the frequency of whole blood donation (INTERVAL): a randomised trial of 45 000 donors. The Lancet 390, 2360–2371 (2017).

35. Karczewski, K. J. et al. The mutational constraint spectrum quantified from variation in 141,456 humans. Nature 581, 434–443 (2020).

36. Minatogawa, M. et al. Expanding the phenotypic spectrum of cardiospondylocarpofacial syndrome: From a detailed clinical and radiological observation of a boy with a novel missense variant in MAP3K7. Am. J. Med. Genet. A 188, 350–356 (2022).

37. Foreman, J. et al. DECIPHER: Supporting the interpretation and sharing of rare disease phenotype-linked variant data to advance diagnosis and research. Hum. Mutat. 43, (2022).

38. Micale, L. et al. Insights into the molecular pathogenesis of cardiospondylocarpofacial syndrome: MAP3K7 c.737-7A>G variant alters the TGFβ-mediated α-SMA cytoskeleton assembly and autophagy. Biochimica et Biophysica Acta (BBA) - Molecular Basis of Disease 1866, 165742 (2020).

39. Morlino, S. et al. A novel MAP3K7 splice mutation causes cardiospondylocarpofacial syndrome with features of hereditary connective tissue disorder. European Journal of Human Genetics 26, 582–586 (2018).

40. Besse, A. et al. TAK1-dependent Signaling Requires Functional Interaction with TAB2/TAB3. Journal of Biological Chemistry 282, 3918–3928 (2007).

41. Aashaq, S., Batool, A. & Andrabi, K. I. TAK1 mediates convergence of cellular signals for death and survival. Apoptosis 24, 3–20 (2019).

42. Cheng, A. et al. 6q25.1 (TAB2) microdeletion syndrome: Congenital heart defects and cardiomyopathy. Am. J. Med. Genet. A 173, 1848–1857 (2017).

43. Engwerda, A. et al. TAB2 deletions and variants cause a highly recognisable syndrome with mitral valve disease, cardiomyopathy, short stature and hypermobility. European Journal of Human Genetics 29, 1669–1676 (2021).

44. Vasilescu, C. et al. Genetic Basis of Severe Childhood-Onset Cardiomyopathies. J. Am. Coll. Cardiol. 72, 2324–2338 (2018).

45. Yin, H. et al. TAB2 deficiency induces dilated cardiomyopathy by promoting RIPK1-dependent apoptosis and necroptosis. Journal of Clinical Investigation 132, (2022).

46. Martin, A. R. et al. PanelApp crowdsources expert knowledge to establish consensus diagnostic gene panels. Nat. Genet. 51, 1560–1565 (2019).

47. Ceyhan-Birsoy, O., Miatkowski, M. M., Hynes, E., Funke, B. H. & Mason-Suares, H. NGS testing for cardiomyopathy: Utility of adding RASopathy-associated genes. Hum. Mutat. 39, 954–958 (2018).

48. Nyuzuki, H. et al. A severe case of cardiospondylocarpofacial syndrome with a novel MAP3K7 variant. Hum. Genome Var. 11, 8 (2024).

